# WHAT IS THE IMPACT OF SELF-EFFICACY, DEPRESSION, ANXIETY, BELIEFS AND PERCEPTION, AND SLEEP QUALITY ON PAIN, DISABILITY, AND QUALITY OF LIFE IN PATIENTS WITH FS? THE PERSPECTIVE FS-PSY-II COHORT STUDY

**DOI:** 10.1101/2025.07.08.25330894

**Authors:** F Brindisino, M Matrisciano, D Feller, E Silvestri, A Fioretti, G Girardi, P Barone, G Merolla, G Guerra

## Abstract

**Background:** Frozen shoulder (FS) is a painful and disabling condition that significantly impairs daily activities, sleep, and quality of life.^1–5^ Beyond structural limitations, FS also involves psychological and behavioural factors such as altered pain beliefs, anxiety, and mood changes.^6,7^ Traditional biomedical models, which separate physical from psychological aspects of illness, often fail to address persistent pain effectively.^8^ FS is not exclusively linked to tissue pathology, as emotional and cognitive components also contribute to symptom intensity. ^8,9^ Despite increasing awareness of the biopsychosocial nature of FS among specialists, these perspectives are not consistently applied in general practice.^10^ However, the broader psychosocial dimensions of FS remain underrepresented in clinical research.^11^

**Aim:** This study aims to investigate whether psychological variables, including self-efficacy, depression, anxiety, pain beliefs, and sleep quality, affect pain, disability, and quality of life after 3 months in patients with FS.

**Methods:** This prospective cohort study will follow the STROBE guidelines. Adults aged 40–65 years diagnosed with primary FS will be recruited. Inclusion criteria include ≥50% restriction in external rotation compared to the unaffected shoulder, and <25% restriction in at least two other planes. Exclusion criteria include prior shoulder surgery, serious comorbidities, or psychiatric conditions interfering with participation. Conservative treatment will include corticosteroid injections, mobilization, stretching, and therapeutic exercise. Patient-reported outcome measures (PROMs) (shoulder Pain and Disability Index - SPADI for shoulder-specific disability, Numeric Rating Scale - NRS for daily and nighttime pain, EQ-5D-5L for quality of life assessment) and psychological variables will be administered at baseline and at 3 months using validated PROMs for self-efficacy, depression, anxiety, pain beliefs, and sleep quality.Primary analysis will examine the relationship between baseline psychological variables and improvement in SPADI using multivariable regression, adjusting for gender and comorbidities. Descriptive statistics, normality tests, and multiple imputation will be used as appropriate. Analyses will be performed with R.

**Discussion and Conclusion:** This study will provide new evidence on how psychological factors influence FS recovery. The findings may support a more integrated and patient-centred approach to treatment, promoting both physical and psychological rehabilitation strategies.

**Study Objective(s) Primary:** To determine if improvement disability (via SPADI), daily and nighttime pain (through NRS) and Quality of life (via EUROQoL 5D-5L), at 3 months is influenced by self-efficacy, depression, anxiety, beliefs and perception, sleep quality in FS patients.

**Study Design** Prospective cohort study

**Subject Population key criteria for Inclusion and Exclusion Inclusion Criteria**

1. adult population (≥ 40 < 65 years old) with FS.FS is defined as range of motion restriction in external rotation at arm by side on the affected limb > 50% with respect to the contralateral limb, restriction <25% in two other planes at least. Moreover, symptoms must remain stable or worsted in the last one month ^2^
2. able to understand and speak Italian
3. have no contraindication for the use of corticosteroids

**Exclusion Criteria**

1. patients with shoulder fractures during the last year
2. patients with rotator cuff repair in the previous year
3. patients with shoulder surgery procedures during the last year
4. patients with shoulder dislocation during the previous year
5. patients with actual serious specific shoulder disorders (i.e., tumour, infection)
6. patients with actual severe psychiatric diagnosed disorders that prevent study participation

**Study Duration**

Each subject’s participation will last up to 3 months; the whole study will be of 12 months.

**Study Phases Screening**

Screening for eligibility: the patients will be screened for inclusion and exclusion criteria and will be informed about the study procedures and aims. All patients will sign a written informed consent to the study.

**Study Treatment Follow-Up**

The treatment will be the best choice up to date highlighted in the literature (corticosteroid infiltrations, mobilization, stretching, exercises)^2, 12–14^ and no added intervention will be administered.

Different study’ time points will be as shown in Table 1 and at the end of the treatment at 3 months, the interview will be conducted online via questionnaire completion

**Efficacy Evaluations**

Primary evaluation measurements will be used to assess the change in disability, pain and quality of life at 3 months. Evidence showed that an important improve in clinical variables is gained in about 3 months from the start of the treatment and continued until 1 year ^15–16^

## Section A - DEMOGRAPHIC INFORMATION REQUESTED

**TABLE 1:**
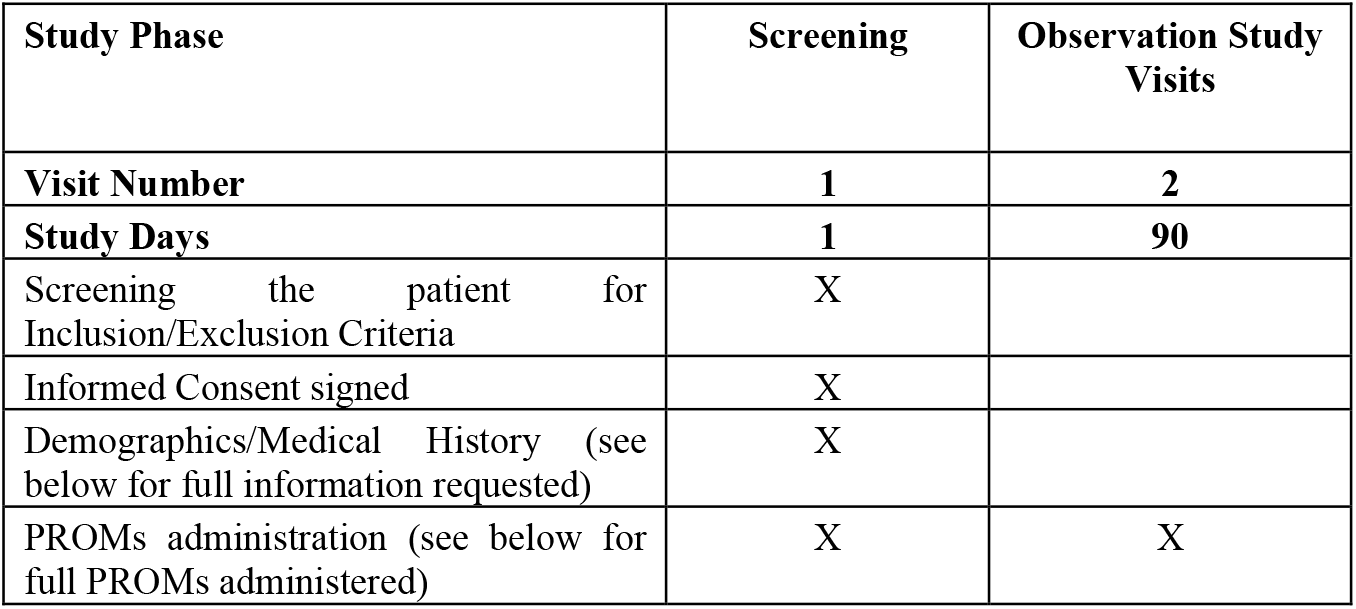
SCHEDULE OF STUDY PROCEDURES.

| Study Phase | Screening | Observation Study Visits |
| --- | --- | --- |
| **Visit Number** | **1** | **2** |
| **Study Days** | **1** | **90** |
| Screening the patient for Inclusion/Exclusion Criteria | X |  |
| Informed Consent signed | X |  |
| Demographics/Medical History (see below for full information requested) | X |  |
| PROMs administration (see below for full PROMs administered) | X | X |

Age (Number)

Gender M□ F□

Work activity

□mainly dynamic

□ mainly static

Education:

Total number of years of education

Physical activity YES□ NO□

Physical activity with the affected shoulder YES□ NO□

Dominant arm Right □ Left□

Affected arm Right □ Left□

Did you suffer from FS to the contralateral side? YES □ NO□

Comorbidities:

DIABETES, YES □ NO □

HYPERGLYCEMIA, YES □ NO □

THYROID DISEASES, YES □ NO □

DUPUYTREN’S DISEASE, YES □ NO □

CARDIOPULMONARY DISEASES, YES □ NO □

RHEUMATOLOGICAL DISEASES, YES □ NO □

METABOLIC DISEASES, YES □ NO □

AUTOIMMUNE DISEASES, YES □ NO □

TUMOR DISEASES (in general), YES □ NO □

NEUROLOGICAL DISEASES, YES □ NO □

NONE

Drugs intake in the last months:

Anxiolytics/Antidepressants YES □ NO □

Barbiturates YES □ NO □

Painkillers YES □ NO □

Anti-inflammatory drugs YES □ NO □

Muscle relaxants YES □ NO □

Corticosteroids YES □ NO □

NONE

### Assessment of Pain and Stiffness

How long have you been suffering from pain by frozen shoulder?

number of WEEKS

How long have you been suffering from stiffness by frozen shoulder?

number of WEEKS

We would like to know how much DAYTIME pain you have experienced over the past week due to your current shoulder problem. Please mark on the line below the level of DAYTIME pain you have experienced in the past week.The horizontal line below represents a scale where the left end indicates no pain at all (0), and the right end indicates the worst pain ever experienced (10).

Please place a mark on the number that best represents the intensity of DAYTIME pain you have felt in your shoulder over the past week.

0 1 2 3 4 5 6 7 8 9 10

We would like to know how much NIGHTTIME pain you have experienced over the past week due to your current shoulder problem. Please mark on the line below the level of NIGHTTIME pain you have experienced in the past week.The horizontal line below represents a scale where the left end indicates no pain at all (0), and the right end indicates the worst pain ever experienced (10).

Please place a mark on the number that best represents the intensity of NIGHTTIME pain you have felt in your shoulder over the past week.

0 1 2 3 4 5 6 7 8 9 10

We would like to know how much stiffness in your shoulder you have experienced over the past week. Please mark on the line below the level of stiffness you have experienced in the past week.

The horizontal line below represents a scale where the left end indicates no stiffness at all (0), and the right end indicates the worst imaginable stiffness (10).

Please place a mark on the number that best represents the feeling of shoulder stiffness you have experienced over the past week.

0 1 2 3 4 5 6 7 8 9 10

## Section B - PROMs ADMINISTERED (outcome)

SPADI total (for pain and disability)

EURO-QoL 5D 5L

## Section C - PROMs ADMINISTERED (exposure)

Pain Self Efficacy Questionnaire (PSEQ)

Beck Depression Inventory 2 (BDI II)

STAI y1; STAI y2

Pain Beliefs and Perception Inventory (PBAPI)

Pittsburgh Sleep Quality Index (PSQI)

### PROMs DESCRIPTION

## Section B

### Shoulder Pain and Disability Index – SPADI- (Giannotta et al., 2024)

The SPADI was developed to measure the pain and disability associated with shoulder pathology. The SPADI is a self-administered index consisting of 13 items divided into two subscales: pain and disability; a 5-item subscale that measures pain and an 8-item subscale that measures disability

### EURO-QoL 5D 5L *(2009 EuroQol Group EQ-5D)*

The 5-level EQ-5D version (EQ-5D-5L) was introduced by the EuroQol Group in 2009 to improve the instrument’s sensitivity and to reduce ceiling effects, as compared to the EQ-5D-3L. The EQ-5D-5L essentially consists of 2 pages: the EQ-5D descriptive system and the EQ visual analogue scale (EQ VAS). The descriptive system comprises five dimensions: mobility, self-care, usual activities, pain/discomfort and anxiety/depression. Each dimension has 5 levels: no problems, slight problems, moderate problems, severe problems and extreme problems. The patient is asked to indicate his/her health state by ticking the box next to the most appropriate statement in each of the five dimensions.

The EQ VAS records the patient’s self-rated health on a vertical VAS, where the endpoints are labelled ‘The best health you can imagine’ and ‘The worst health you can imagine’. The VAS can be used as a quantitative measure of health outcome that reflect the patient’s own judgement.

## Section C

### Self-Efficacy (Pain Self Efficacy Questionnaire – PSEQ) (Chiarotto, 2015)

The PSEQ is used to measure pain self-efficacy and consists of 10 items representing different daily activities (e.g., I can do most of the household chores) or general aspects of life (e.g., I can still accomplish most of my goals in life). For each item, the patient must rate how confident he or she feels to perform these activities, despite the presence of pain. Items are rated on a 7-point Likert scale ranging from 0 (not at all confident) to 6 (completely confident). The total score of the questionnaire can range from 0 to 60, with higher scores indicating higher pain self-efficacy.

### Beck Depression Inventory II (Sica, 2007)

Beck Depression Inventory II Edition (BDI-2) is a 21-item multiple-choice self-report inventory for assessing affective-somatic (AS) and cognitive dimensions (C) of depression severity. The total score ranges from 0 to 63; in particular, a score from 10 to 18 indicates a mild to moderate depression, a score from 19 to 29 indicates a moderate to severe depression, and a score higher than 30 is indicative of a severe depression level.

### STAI Y1Y2 (Ilardi, 2021)

State-Trait Anxiety Inventory (STAI Form Y1 and Y2) is a 40-item multiple-choice self-report inventory divided into two sub-tests, each having 20 items (for both state- and trait-anxiety) aimed at assessing and quantifying anxiety disorder in adults. All items are rated on a 4-point Likert scale. The recommended clinical cut-off is >46.

### Pain Beliefs and Perceptions Inventory (PBAPI) (Monticone, 2013)

This is a 16-item questionnaire, and patients rate their beliefs using a 4-point Likert scale ranging from −2 (total disagreement) to −2 (total agreement); item Nos. 3, 9, 12 and 15 are reverse scored. For each subscale, the scores of the responses to the items that are answered are added and divided by the number of items answered, higher scores indicate greater endorsement of the beliefs.

### Pittsburgh Sleep Quality Index (PSQI) (Curcio, 2013)

The 19-item Pittsburgh Sleep Quality Index (PSQI) is probably the most used retrospective self-report questionnaire, that measures sleep quality over the previous month. Seven clinically derived domains of sleep difficulties (sleep quality, sleep latency, sleep duration, habitual sleep efficiency, sleep disturbances, use of sleeping medications, and daytime dysfunction) are assessed by the questionnaire. Taken together, these sleep domains are scored as a single factor of global sleep quality. Usually, a global score higher than 5 is considered as an indicator of relevant sleep disturbances in at least two components or of moderate difficulties in more than three components. More recently, an overlapping of some components has been observed, and three distinct factors have been extracted: sleep efficiency (including sleep duration and habitual sleep efficiency), perceived sleep quality (including subjective sleep quality, sleep latency and use of sleeping medication), and daily disturbances (including sleep disturbances and daytime dysfunction)

## Compliance Statement

This study will be reported in accordance with the STROBE reporting guideline.

## 1. General Schema of Study Design

prospective cohort study

### 1.1 Study Duration, Enrolment and Number of Sites

#### 1.1.1 Duration of Study Participation

The study duration per subject will be up to 90 days, with follow-up divided as showed in Table 1.

## 2 STUDY PROCEDURES

1. identification of potentially eligible subjects
2. screening for inclusion and exclusion criteria
3. willing to participate and informed consent signed
4. collections of demographic variables, administration of PROMs for exposition and outcome
5. treatment
6. evaluations at follow up points

### 2.1 Screening Visit (*Baseline*)

- Screening for inclusion and exclusion criteria
- Informed Consent
- Collection of demographic variables using Google Forms
- PROMs administration using Google Forms (https://docs.google.com/forms/u/0/)

### 2.2 Treatment Period

Pertaining to the observed improvements in FS patients, two recent studies have demonstrated a more pronounced improvement occurring within the initial three months of treatment. This finding led us to establish the three-month mark for our follow-up assessment.

#### 2.2.1 Visit 2 (90 days)

1. PROMs administration through Google Form

## STATSTICAL ANALYSIS

The shape of the distribution of the variables related to the population will be evaluated through a graphical analysis.

The collected variables will be described using descriptive statistics. In particular, mean and standard deviation, median with the interquartile range (IQR), and relative frequency and percentages will be calculated for the variables with normal, non-normal and categorical distribution, respectively.

To describe the changes in clinical and functional variables between admission and discharge, graphical analyses will be used. Also, we will report descriptive statistics at the various follow-ups (**Table 1**).

Lost at follow-up and dropouts will be counted and evaluated. Missing data (referable to exposure and outcome variables) will be imputed through a “multiple imputation” procedure. We will impute a number of datasets equal to the number of subjects with at least one missing data. The reasons of drop out will be presented if the research team is aware of them. Patients who withdraw their informed consent during follow-up will not be included in the analyses. Baseline characteristics of patients lost to follow-up will be compared with those remaining in the study.

## PRIMARY ANALYSIS

Depression, anxiety (via STAY1 and STAY2), self-efficacy, beliefs and perceptions, and sleep quality will be evaluated as potential prognostic factors in a multivariable linear regression model. The dependent variables will be daytime and nighttime pain, disability, and quality of life (each treated as continuous outcomes). The model will be adjusted for gender and for the presence of comorbidities (diabetes and/or thyroid disorders).

The assumption of absence of multicollinearity will be assessed through the variance inflation factor (VIF) (with a threshold > 5). The possible presence of influential values will be examined using Cook’s distance, applying a threshold of 4/n. The normality of the residuals will be assessed through visual inspection of Q–Q plots.

The required sample size has been estimated as 80 patients, based on the rule of thumb of at least 10 patients per coefficient included in the regression model (i.e., 8 coefficients).

All statistical analyses will be performed using R.

### 2.3 Subject Completion/Withdrawal

Subjects may withdraw from the study at any time without prejudice to their care. They may also be discontinued from the study at the discretion of the Investigator for lack of adherence to study treatment or visit schedules, adverse events, or any other reason (to be documented). The Investigator may also withdraw subjects who violate the study plan, or to protect the subject for reasons of safety or administrative reasons. It will be documented whether each subject completes the clinical study. If the Investigator becomes aware of any serious, related adverse events after the subject completes or withdraws from the study, he will record it.

### 2.4 Clinical Adverse Events

Clinical adverse events (AEs) will be monitored throughout the study and assessed at 3 months.

### 2.5 Definition of an Adverse Event

An AE is any untoward medical occurrence in a subject who has received an intervention (drug, biological, or other intervention). The occurrence does not necessarily have a causal relationship with the treatment. An AE can therefore be any unfavourable or unintended sign (including an abnormal laboratory finding, for example), symptom, or disease temporally associated with using a medicinal product, whether considered related to the medicinal product.

All AEs (including serious AEs) will be noted in the study records and on the case report form with a complete description including the nature, date and time of onset, determination of non-serious versus serious, intensity (mild, moderate, severe), duration, causality, and outcome of the event.

## Data Availability

All data produced in the present study are available upon reasonable request to the authors

